# Retrospective cohort analysis of antiretroviral therapy initiation timelines and clinical outcomes in adults with HIV and TB disease in KwaZulu-Natal, South Africa

**DOI:** 10.1101/2025.03.05.25323306

**Authors:** Claudia J Jansen van Vuuren, Johan van der Molen, Yukteshwar Sookrajh, Thulani Ngwenya, Thokozani Khubone, Siyabonga Mkhize, Kwabena Asare, Kogieleum Naidoo, Richard Lessells, Lara Lewis, Nigel Garrett, Jienchi Dorward

## Abstract

**Background:** We aimed to determine antiretroviral therapy (ART) initiation timing and outcomes in people living with HIV (PLHIV) receiving tuberculosis treatment in KwaZulu-Natal, South Africa.

**Methods:** We performed a retrospective cohort analysis of routinely collected de-identified data from 62 clinics including PLHIV not already receiving ART aged ≥16 years, starting tuberculosis treatment between October 2016–November 2019. Multivariable Poisson regression models with robust standard errors evaluated associations between timing of ART initiation (after starting tuberculosis treatment) and successful tuberculosis treatment, and 6-month HIV viral load (VL) <50 copies/mL.

**Results:** Among 5,548 PLHIV with tuberculosis, 29.8% initiated ART within 15 days (“early”), 36.2% in 16-56 days, 8.7% in 57-210 days, with 25.3% not initiating ART by 7 months. Proportions with successful tuberculosis treatment were similar comparing 16-56 and 57-210 days to early initiation, with a lower likelihood of successful tuberculosis outcome with no ART within 7 months (adjusted risk ratio [aRR] 0.81 [0.77-0.86], p<0.001). In those with a known VL 6 months post-ART initiation (n=2,658), initiation within 57-210 days had a lower likelihood of viral suppression (aRR 0.90 [0.82-0.99], p<0.03).

**Conclusion:** Although <30% of PLHIV with tuberculosis initiated ART early, this was associated with better tuberculosis outcomes and VL suppression.

## Introduction

Tuberculosis (TB) disease remains a global public health challenge, with an estimated 10.6 million new TB diagnoses in 2022.^1^ In South Africa (SA), TB was the second leading cause of deaths in 2021,^2^ with an estimated prevalence amongst all adults, and those living with HIV, of 852 and 1,734 per 100,000 population, respectively.^3^

Untreated HIV infection increases the risk of TB,^4^ and management of HIV/TB coinfection is complicated by increased pill burden, drug interactions, drug-induced liver injury and paradoxical TB-immune reconstitution inflammatory syndrome (IRIS).^5, 6^ SA has the highest HIV incidence globally, and KwaZulu-Natal is the worst affected province, with an HIV prevalence of 27%^7^, where HIV and TB infections remain the leading causes of death in people aged 15-64 years.^8^

Initiation of antiretroviral therapy (ART) during TB therapy in HIV/TB has been shown in multiple trials to have mortality benefit,^9–12^ with early ART in those with CD4 count below 50 cells/µL conveying a higher risk of IRIS but improved survival, as well as a lower risk of TB treatment failure.^13–15^ Early ART initiation offers further potential benefits, e.g. viral suppression and retention-in-care are improved by early ART initiation in PLHIV without TB. However, the effects of early initiation on these outcomes in PLHIV with TB disease are not conclusively known.^16, 17^

The World Health Organization (WHO) has recommended that “ART should be started as soon as possible within two weeks of initiating TB treatment, regardless of CD4 count, among people living with HIV.”^18^ The South African National Department of Health (NDOH), which provides fully integrated HIV and TB care free at the point of service, advises commencing ART within 2 weeks of starting TB treatment in those with CD4 count below 50 cells/µL, but after 8 weeks in those with CD4 count above this threshold,^19^ while the Southern African HIV Clinicians Society advises delaying ART in this group “until 8 weeks…but no later”.^20^

Considering these different recommendations, we aimed to determine the timepoints at which patients are initiated on ART after starting TB treatment in the public health setting in KwaZulu-Natal, SA; and to evaluate subsequent TB outcomes and HIV viral load (VL) suppression.

## Methods

### Study design, setting and data sources

We performed a retrospective cohort analysis using routine, de-identified data in the TIER.Net database, an electronic register used in the South African public sector recording demographic, clinical and clinic visit data for HIV care^21^. We used data from 56 urban clinics run by the eThekwini Municipality Health Unit, in the largest urban centre in KwaZulu-Natal, and six rural clinics run by Bethesda Hospital, in the largely rural uMkhanyakude District of northern KwaZulu-Natal. Integrated HIV and TB care is provided free-of-charge at these clinics in accordance with South African guidelines.^22^

### Participants

We included PLHIV starting TB treatment between 1 Oct 2016 and 1 November 2019, aged ≥16 years, and who had not yet initiated ART. We analysed data for patients up until the end of their TB treatment and excluded patients who had no documented end date of TB treatment by the data cut-off date (01 June 2020). For patients who had more than one recorded TB episode during the study period, only the first was analysed. There were no records of drug-resistant TB (DR-TB) in the cohort analysed.

### Variables

We grouped records into three TB outcome categories which correspond with the South African National Department of Health definitions, and WHO TB treatment outcome definitions of 2013.^23^ We considered those who completed TB treatment or were cured based on sputum results as having “successful” TB outcomes; those who failed treatment, defaulted treatment, or died as having “unsuccessful” TB outcomes; and in those patients where there was no treatment outcome assigned in the records, we designated their TB outcome as being “unknown”. For assessing patients’ TB outcome, those with unknown outcomes were excluded from the analysis, to arrive at the primary outcome of successful vs unsuccessful TB treatment. A second analysis was conducted to investigate the outcome of HIV viral suppression <50 copies/mL 6 months after ART initiation (this being the recommended timepoint for VL measurement per South African HIV guidelines). Because VLs are not always taken according to the recommended schedule, we allowed a window of 90-270 days and used the VL closest to 180 days.

The primary exposure of interest for both analyses was the time to ART initiation following commencement of TB treatment, grouped into ≤15, 16-56, 57-210 or ≥210 days to align to the timeframes in the different ART guidelines. We used 15 days rather than 14 days as the cut-off for “2 weeks” due to possible variation in clinicians’ practices of assigning the first day of ART as “Day 0” or “Day 1”. We chose a value of 210 days after examining the observed distribution of TB treatment durations in the dataset. Potential confounding variables included demographics (gender, age group and rural vs urban district); baseline clinical characteristics (pregnancy, CD4 count, site of TB disease) and treatment-related factors (TB episode type, e.g. newly diagnosed, recurrence or relapse). Because CD4 counts are not always taken to schedule, we allowed a window of 180 days before until 60 days after TB treatment start and used the record closest to the TB treatment starting date.

### Statistical analysis

Frequencies and percentages were used to summarize the cohort characteristics and to assess missing data. We conducted univariable and multivariable Poisson regression models with robust standard errors to evaluate the association between time to ART initiation, and TB and HIV suppression outcomes, adjusted for potential confounding variables. We used robust standard errors to account for clustering by facility. Potential confounders that were included in the multivariable model were selected based on the data available and clinical plausibility. Pregnancy status and district were excluded as covariates from the analyses due to small sample sizes. In the primary analysis, where data was missing for potential confounding variables, this was included in the analysis as a separate category (“Missing”).

We determined, a priori, to assess whether any effect of time to ART initiation was modified by baseline CD4 count by conducting a sensitivity analysis with an interaction term between CD4 count and time to ART initiation, and adjusted for potential confounders. In this analysis, those with missing data in exposure and/or outcome were excluded. We also conducted sensitivity analyses where those with unknown TB outcomes were classified as ‘unsuccessful’, and also excluding people with extra-pulmonary TB who may have been more likely to be diagnosed in secondary care which could have lengthened time to being seen for ART in primary care.

Statistical analysis was performed using R 4.2.1 (R Foundation for Statistical Computing, Vienna, Austria).

### Ethical considerations

This study was approved by the University of Kwazulu-Natal Biomedical Research Ethics Committee (BE646/17), the eThekwini Municipality Health Unit, the Bethesda Hospital Ethics Committee and the KwaZulu-Natal Department of Health Provincial Research Ethics Committee. The need for informed consent was waived as de-identified, routinely collected data was analysed.

## Results

A total of 20,063 people had a TB episode in the period described of whom 5,548 met the inclusion criteria (Fig. 1). The median age was 35 years, and more people (62.0%, n=3,439) were male (Table 1). The baseline CD4 count was <50 cells/µL in 11.5% (n=640) and unknown in 44.9% (n=2,492). Pulmonary TB was most common (87.1%, n=4,833) and for 91.1% of people (n=5,055) it was their first episode of TB disease. 192/5,548 (3.5%) had more than one episode of TB in the study period. Drug-sensitive TB regimens were commenced in 5,544 people (99.9%), and 4 people had no specified regimen.

**Figure 1:**
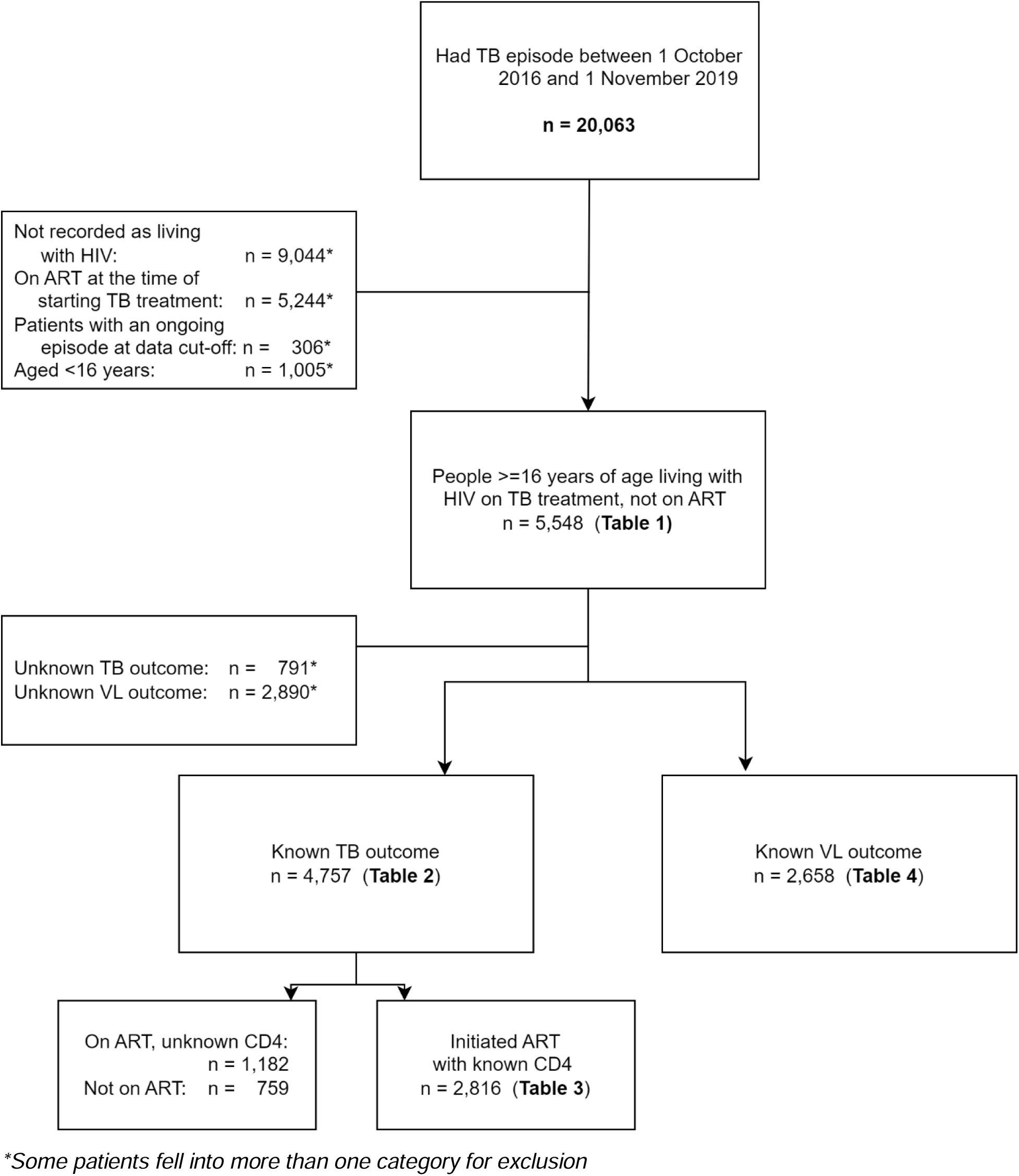
CONSORT diagram for inclusion of data in analysis. Records containing routinely collected clinical data were reviewed for patients with TB attending 61 clinics in KwaZulu-Natal between October 2016 and November 2019 (n = 20,063).

**Table 1:** Variables and outcomes of patients commencing ART after TB initiation (N=5,548)

| Variable | Group | n/N<br>(% of total) | Outcome |  |  |
| --- | --- | --- | --- | --- | --- |
|  |  |  | Successful<br>n (% of group) | Unsuccessful<br>n (% of group) | Unknown<br>n (% of group) |
|  |  | **5,548 (100.0)** | **3,937 (71.0)** | **820 (14.8)** | **791 (14.3)** |
| **Time to ART initiation (days)** | ≤15 | 1,652 (29.8) | 1,343 (81.3) | 188 (11.4) | 121 (7.3) |
|  | 16-56 | 2,011 (36.2) | 1,577 (78.4) | 267 (13.3) | 167 (8.3) |
|  | 57-210 | 481 (8.7) | 387 (80.5) | 69 (14.3) | 25 (5.2) |
|  | Not initiated within 7 months | 1,404 (25.3) | 630 (44.9) | 296 (21.1) | 478 (34.0) |
| **Gender** | Male | 3,439 (62.0) | 2,388 (69.4) | 565 (16.4) | 486 (14.1) |
|  | Female | 2,109 (38.0) | 1,549 (73.4) | 255 (12.1) | 305 (14.5) |
| **Age group** | 16-25 | 580 (10.5) | 393 (67.8) | 100 (17.2) | 87 (15.0) |
|  | 26-35 | 2,305 (41.5) | 1,645 (71.4) | 347 (15.1) | 313 (13.6) |
|  | 36-45 | 1,739 (31.3) | 1,242 (71.4) | 241 (13.9) | 256 (14.7) |
|  | 46-55 | 699 (12.6) | 506 (72.4) | 91 (13.0) | 102 (14.6) |
|  | >55 | 225 (4.1) | 151 (67.1) | 41 (18.2) | 33 (14.7) |
| **Pregnancy status at ART initiation** | Not pregnant | 2,016 (36.3) | 1,476 (73.2) | 245 (12.2) | 295 (14.6) |
|  | Pregnant | 47 (0.8) | 35 (74.5) | 6 (12.8) | 6 (12.8) |
|  | Not applicable | 46 (0.8) | 38 (82.6) | 4 (8.7) | 4 (8.7) |
|  | Unknown | 3,439 (62.0) | 2,388 (69.4) | 565 (16.4) | 486 (14.1) |
| **CD4 count (cells/μL)** | <50 | 640 (11.5) | 505 (78.9) | 72 (11.2) | 63 (9.8) |
|  | 50-199 | 1,217 (21.9) | 990 (81.3) | 136 (11.2) | 91 (7.5) |
|  | 200-349 | 647 (11.7) | 540 (83.5) | 65 (10.0) | 42 (6.5) |
|  | 350-499 | 296 (5.3) | 234 (79.1) | 41 (13.9) | 21 (7.1) |
|  | ≥500 | 256 (4.6) | 196 (76.6) | 43 (16.8) | 17 (6.6) |
|  | Missing | 2,492 (44.9) | 1,472 (59.1) | 463 (18.6) | 557 (22.4) |
| **District** | Rural | 281 (5.1) | 205 (73.0) | 30 (10.7) | 46 (16.4) |
|  | Urban | 5,267 (94.9) | 3,732 (70.9) | 790 (15.0) | 745 (14.1) |
| **TB category** | Newly diagnosed | 5,055 (91.1) | 3,610 (71.4) | 728 (14.4) | 717 (14.2) |
|  | After relapse | 407 (7.3) | 269 (66.1) | 68 (16.7) | 70 (17.2) |
|  | After default | 45 (0.8) | 29 (64.4) | 14 (31.1) | 2 (4.4) |
|  | After failure | 20 (0.4) | 11 (55.0) | 8 (40.0) | 1 (5.0) |
|  | Unknown | 21 (0.4) | 18 (85.7) | 2 (9.5) | 1 (4.8) |
| **TB site** | Pulmonary TB | 4,833 (87.1) | 3,445 (71.3) | 709 (14.7) | 679 (14.0) |
|  | Extra-pulmonary TB | 436 (7.9) | 318 (72.9) | 62 (14.2) | 56 (12.8) |
|  | Nervous system TB | 111 (2.0) | 80 (72.1) | 17 (15.3) | 14 (12.6) |
|  | Unknown | 168 (3.0) | 94 (56.0) | 32 (19.0) | 42 (25.0) |

Note: TB outcomes were defined as follows: Successful – those who completed TB treatment or were cured based on sputum results. Unsuccessful – those who failed treatment, defaulted treatment, or died.

ART was initiated within 15 days of commencing TB treatment in 29.8% (n=1,652) of people, 16-56 days in 36.2% (n=2,011), 57-210 days in 8.7% (n=481); while in 25.3% (n=1,404) of people, ART was initiated after 7 months, or not at all. ART was initiated on the same day as starting TB treatment in 100 people (1.8%). Of those who initiated ART within 7 months, 4,052 of 4,144 (97.8%) initiated on tenofovir disoproxil fumarate, emtricitabine and efavirenz.

### TB outcomes

Of the 5,548 people meeting inclusion criteria, 2,957 (53.3%) completed TB treatment, 980 (17.7%) were cured, 596 (10.7%) defaulted treatment, 200 (3.6%) died, 24 (0.4%) failed treatment, and 791 (14.3%) were lost to follow up. Of the 100 people who initiated ART and TB on the same day, 76 (76%) completed treatment or were cured, 15 (15%) failed/defaulted/died, and 9 (9%) were lost to follow up.

Considering only those with known TB outcomes (n=4,757, Fig. 1) and after adjusting for potential confounders, people who did not initiate ART within 7 months had a 19% lower risk of successful outcomes (adjusted risk ratio [aRR] 0.81 [0.77-0.86], p<0.001) compared to those initiating within 15 days (Table 2). There was no difference in outcomes when comparing groups initiating ART at other timepoints to the group initiating within 15 days. A sensitivity analysis including the 791 people who had no treatment outcome assigned in their records (which were excluded as having “unknown” outcomes in the primary analysis) in the “unsuccessful” category produced similar results (Supplementary Table S1), as did an analysis of TB outcomes which included only people diagnosed with pulmonary TB and known outcomes (n=4,154, Supplementary Table S2).

**Table 2:**
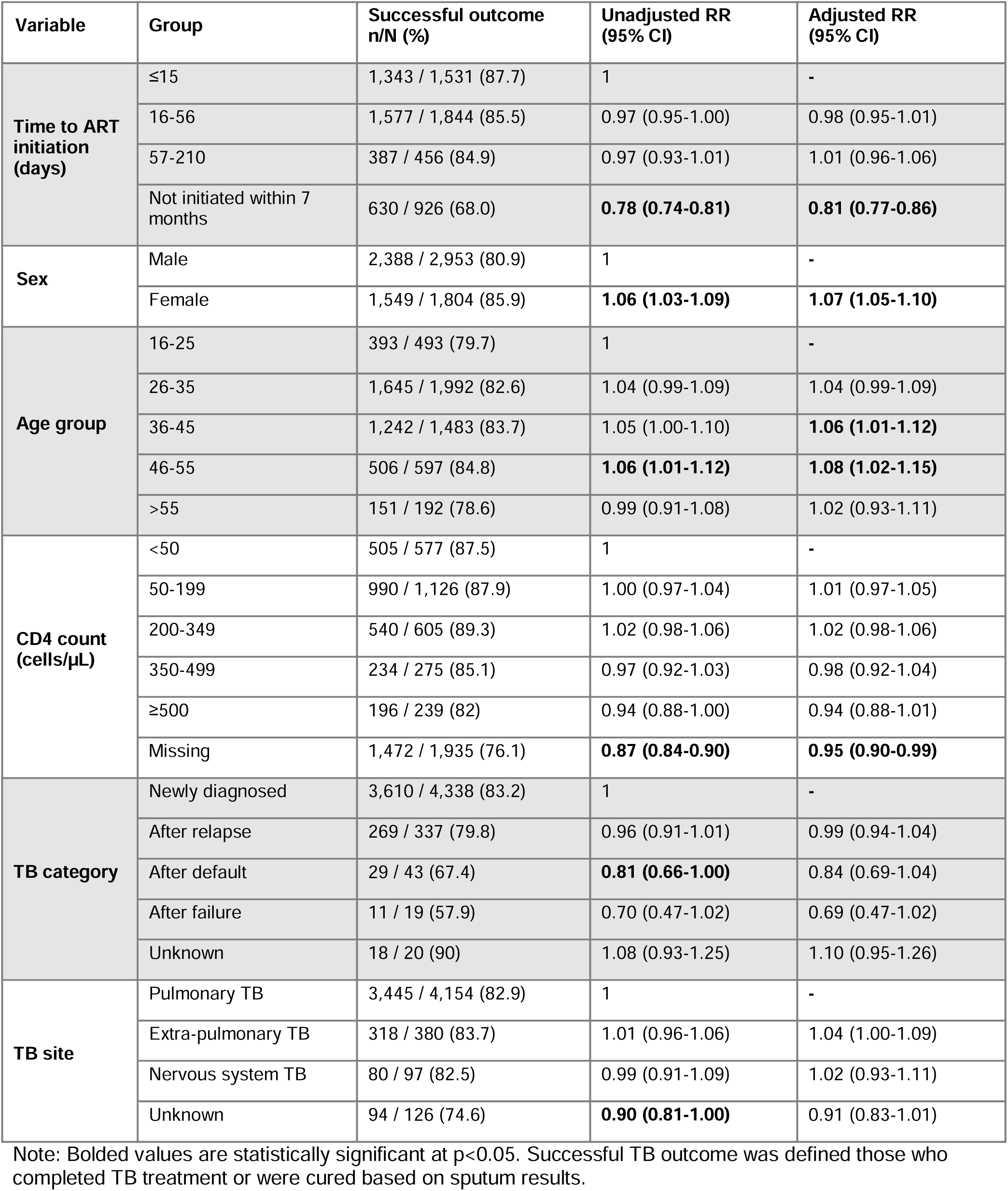
Univariable and multivariable Poisson regression models of factors associated with successful TB outcomes, among people living with HIV on TB treatment (N=4,757)

We conducted a sensitivity analysis with an interaction term between CD4 count and time to ART initiation (Table 3). We included only people with a known baseline CD4 count and those who initiated ART (n=2,816, see Fig. 1). Among people with CD4 count≥50 cells/µL (n=2,241), a successful TB outcome was less likely when ART was initiated after 16-56 days rather than within two weeks after commencing TB therapy (aRR 0.96 [0.93-0.99], p=0.016), with fewer people being cured (22.4% vs 25.9%, see Supplementary Table S3) and more people failing treatment (0.8% vs 0.4%), defaulting (10.6% vs 8.5%) and dying (2.6% vs 1.6%). Among people with CD4 count <50cells/µL (n=575), the majority had successful TB outcomes, and there were no statistically significant differences in these outcomes between the different initiation timelines, with more detail of outcomes shown in Supplementary Table S3.

**Table 3:** Successful TB outcome by CD4 count and time to ART initiation (N = 2,816)

|  | Successful TB outcomes in patients with CD4 <50 cells/μL |  | Successful TB outcomes in patients with CD4 ≥50 cells/μL |  |
| --- | --- | --- | --- | --- |
| Time to ART initiation (days) | n/N (%) | Adjusted RR (95% CI) | n/N (%) | Adjusted RR (95% CI) |
| ≤15 | 219 / 251 (87.3) | 1 | 906 / 1,012 (89.5) | 1 |
| 16–56 | 270 / 307 (87.9) | 1.01 (0.95-1.07) | 996 / 1,158 (86.0) | **0.96 (0.93-0.99)** |
| 57–210 | 14 / 17 (82.4) | 0.96 (0.77-1.20) | 57 / 71 (80.3) | 0.91 (0.81-1.02) |

Note: Bolded values are statistically significant at $p < 0.05$ . Successful TB outcome was defined those who completed TB treatment or were cured based on sputum results.

Aside from time to ART initiation, additional variables that were associated with successful TB outcomes (Table 2) included female gender (aRR 1.07 [1.05-1.10], p<0.001) and being aged 36-45 years (aRR 1.06 [1.01-1.12], p=0.016), and 46-55 years (aRR 1.08 [1.02-1.15], p=0.006) versus being aged 16-25.

### HIV outcomes

In people with a known HIV VL at 6 months after ART initiation (n=2,658, Table 4), those who initiated ART 57-210 days after commencing TB therapy were less likely to be virally suppressed (182/257, 70.8%, aRR 0.90 [0.82-0.99], p=0.03) compared to ART initiation within 15 days (849/1083, 78.4%). In the group where ART was initiated after 210 days subsequent 6-month viral suppression was similar to the 57-210 day group, (64/91, 70.3%), but the 95% confidence interval of the adjusted risk ratio crossed one (0.77–1.03), likely due to the small numbers in this group. Of the 100 people who initiated ART and TB treatment on the same day, 45 (45%) had a suppressed VL at 6 months, 12 (12%) were not suppressed, and 43 (43%) did not have a VL recorded.

**Table 4:** Univariable and multivariable Poisson regression models of factors associated with viral load suppression, among people living with HIV on TB treatment who initiated ART (N=2,658).

| Variable | Group | Suppressed<br>n/N (%) | Unadjusted RR<br>(95% CI) | Adjusted RR<br>(95% CI) |
| --- | --- | --- | --- | --- |
| **Time to**<br>**ART**<br>**initiation**<br>**(days)** | ≤15 | 849 / 1,083 (78.4) | 1 | - |
|  | 16-56 | 953 / 1,227 (77.7) | 0.99 (0.95-1.03) | 0.99 (0.95-1.04) |
|  | 57-210 | 182 / 257 (70.8) | **0.90 (0.83-0.98)** | **0.90 (0.82-0.99)** |
|  | Not initiated within 7<br>months | 64 / 91 (70.3) | 0.90 (0.78-1.03) | 0.89 (0.77-1.03) |
| **Sex** | Male | 1,257 / 1,636 (76.8) | 1 | - |
|  | Female | 791 / 1,022 (77.4) | 1.01 (0.97-1.05) | 1.02 (0.97-1.06) |
| **Age group** | 16-25 | 190 / 252 (75.4) | 1 | - |
|  | 26-35 | 824 / 1,091 (75.5) | 1.00 (0.93-1.08) | 1.01 (0.93-1.09) |
|  | 36-45 | 685 / 884 (77.5) | 1.03 (0.95-1.11) | 1.04 (0.96-1.13) |
|  | 46-55 | 270 / 337 (80.1) | 1.06 (0.97-1.16) | 1.07 (0.98-1.17) |
|  | >55 | 79 / 94 (84) | 1.11 (1.00-1.25) | **1.13 (1.01-1.27)** |
| **CD4 count**<br>**(cells/μL)** | <50 | 294 / 421 (69.8) | 1 | - |
|  | 50-199 | 607 / 791 (76.7) | **1.10 (1.02-1.18)** | **1.10 (1.02-1.18)** |
|  | 200-349 | 344 / 416 (82.7) | **1.18 (1.10-1.28)** | **1.18 (1.10-1.28)** |
|  | 350-499 | 151 / 186 (81.2) | **1.16 (1.06-1.28)** | **1.16 (1.06-1.28)** |
|  | ≥500 | 137 / 158 (86.7) | **1.24 (1.14-1.36)** | **1.25 (1.14-1.36)** |
|  | Missing | 515 / 686 (75.1) | 1.08 (1.00-1.16) | **1.12 (1.04-1.22)** |
| **TB category** | Newly diagnosed | 1,907 / 2,466 (77.3) | 1 | - |
|  | After relapse | 116 / 159 (73) | 0.94 (0.86-1.04) | 0.94 (0.85-1.03) |
|  | After default | 10 / 13 (76.9) | 0.99 (0.74-1.34) | 1.01 (0.74-1.37) |
|  | After failure | 5 / 8 (62.5) | 0.81 (0.47-1.38) | 0.80 (0.46-1.40) |
|  | Unknown | 10 / 12 (83.3) | 1.08 (0.84-1.39) | 1.10 (0.86-1.40) |
| **TB site** | Pulmonary TB | 1,824 / 2,355 (77.5) | 1 | - |
|  | Extra-pulmonary TB | 148 / 189 (78.3) | 1.01 (0.94-1.09) | 1.02 (0.94-1.10) |
|  | Nervous system TB | 34 / 52 (65.4) | 0.84 (0.69-1.03) | 0.85 (0.70-1.04) |
|  | Unknown | 42 / 62 (67.7) | 0.87 (0.74-1.04) | 0.89 (0.75-1.06) |

Note: Bolded values are statistically significant at p<0.05.

In an analysis including only people diagnosed with pulmonary TB, there were no significant differences in risk of achieving viral suppression comparing different initiation timepoints (Supplementary Table S4).

Other variables associated with better viral suppression included age >55 years (aRR 1.13 [1.01-1.27], p=0.031, compared to 16-25 years; and higher CD4 counts compared to CD4 <50 cells/µL: CD4 50-199 cells/µL (aRR 1.10 [1.02-1.18], p=0.014), 200-349 cells/µL (aRR 1.18 [1.10-1.28], p<0.001, 350-499 cells/µL (aRR 1.16 [1.06-1.28], p=0.002 and ≥500 cells/µL (aRR 1.25 [1.14-1.36], p<0.001). People with missing CD4 count (n=686) were also more likely to achieve viral suppression (aRR 1.12 [1.04-1.22], p=0.004).

## Discussion

While WHO guidelines recommend ART initiation as soon as possible within two weeks of starting (regardless of CD4 count), South African guidelines recommend ART initiation after 8 weeks in people with CD4 >50 cells/µL. In this South African cohort, we found wide variations in the timelines to ART initiation, with only 30% initiating within two weeks of starting TB treatment. Overall, we found worse outcomes in people who did not initiate ART within 7 months, but no difference in TB outcomes between people initiating ART within 15 days, 16-56 days or 57-210 days after starting TB treatment. Those with CD4 count ≥50 cells/µL who initiated ART early appeared to have slightly better TB outcomes compared to those who initiated later.

Almost all people in our dataset were on drug-sensitive TB regimens (apart from a handful of unspecified regimens), as drug-resistant TB is treated at specialised centres, not at primary care level.

The 2021 WHO guidance recommending early ART initiation in the setting of TB disease is based on data from randomised controlled trials which showed, firstly, that mortality is similar when ART is initiated within two weeks versus between two and eight weeks (with moderate-certainty evidence); and, secondly, that mortality is similarly reduced in those with CD4<50 and ≥50 cells/µL (with low-certainty evidence).^24^ Our observational data findings add to this evidence, showing that in this South African cohort, people who had ART initiation within two weeks of starting TB treatment generally had successful outcomes, even in those with higher CD4 counts. People with higher CD4 counts have, in the past, had their ART initiation delayed due to the perceived risk of TB-IRIS, but which does not increase the mortality risk of early initiation, as death from TB-IRIS is uncommon.^24^

Aside from overall mortality benefit, early ART initiation improves viral suppression and retention-in-care in PLHIV without TB, but these effects in PLHIV with TB disease have not conclusively been shown.^16, 17^ In some trials, there was no difference in virological suppression when comparing early versus late initiation of ART in the context of TB,^15, 25^ but other studies suggest that retention-in-care and virological suppression in people with TB symptoms (but low rates of confirmed TB) was improved by same-day initiation.^26^ The 2021 WHO guidance summarizes, with low-certainty evidence, that VL suppression “may not differ” between those initiating ART within two weeks compared to between two and eight weeks,^24^ and the findings presented here also suggest that 6-month viral suppression is more likely when ART is initiated within 8 weeks. Importantly, the VL measurement results in this dataset are representative of people’ VLs 6 months post-ART initiation, which were performed by healthcare workers following the HIV guidelines in SA. We expect this relationship would have been more robust if comparison was possible between VLs at a timepoint relative to baseline (e.g. 6 months after commencing TB treatment), as people who initiated sooner would achieve viral suppression sooner.

Identifying PLHIV with TB symptoms, or confirmed TB, who may benefit from early (or even same-day) initiation remains a difficult clinical decision for providers. In SA, a public health approach to HIV is followed and guidelines should be as clear as possible for the wide range of healthcare workers delivering HIV and TB care.^27^ The groups who had more successful TB outcomes in our study were consistent with previous studies;^28,29^ namely, females and middle-aged adults, in contrast to males, adolescents and the elderly, who had less successful outcomes. Adherence to TB treatment and ART is essential for positive TB and HIV outcomes, and males, adolescents and young adults are known to experience complex barriers to adherence.^29, 30^ Although elderly people had worse TB outcomes than some other age groups, they were more likely to have suppressed VLs six months after ART initiation. Those who started ART with higher CD4 counts were also more likely to reach viral suppression, consistent with improved outcomes among those starting ART earlier during HIV disease.^31, 32^

Our study has several strengths and limitations. We used data from public sector clinics which better reflects real life conditions than clinical trials. However, the routine data in the TIER.net database was not created specifically to answer this research question,^33^ and so other outcomes such as TB-IRIS and adverse drug reactions were not available. Furthermore, errors in data capturing, misclassification of patient outcomes, missing data and poor recording of inter-facility transfers^34^ may influence our results. For example, in our analysis, over a quarter of the people initiating TB treatment in this population did not have a record of initiating ART by the end of their TB treatment, a proportion higher than expected, especially as SA’s integrated HIV and TB services have been shown to improve the chances of PLHIV on TB treatment being initiated on ART.^35^ We used primary care data, where the majority of people receive HIV and TB treatment, but the sickest people who commenced TB treatment in hospital and did not survive to discharge would not have been included. While we adjusted for baseline demographic and clinical variables, unmeasured confounders could include clinician discretion to delay or expedite ART initiation. Finally, most people were receiving efavirenz in our analysis, and so further analyses of outcomes on dolutegravir are warranted.

### Implications for research and policy

Policies to date have been based on findings from several clinical trials, which may not be generalizable to populations with TB initiating ART in routine national programmes. This research suggests that harmonization of the SA guidelines to the WHO guidelines, to initiate ART within 2 weeks of TB treatment regardless of CD4 count, may be of benefit to people in SA for both TB outcome and viral suppression. Further research should consider how early/same-day ART initiation may improve long-term outcomes such as retention-in-care and viral suppression.

## Conclusion

Less than a third of PLHIV with TB disease initiated ART within 15 days of starting TB treatment. Early initiation of ART is associated with better TB outcomes and HIV VL suppression.

## Acknowledgements

We gratefully acknowledge the staff and patients of eThekwini Municipality and Bethesda Hospital primary care clinics.

## Authors’ contributions

CJvV, JD and NG conceived the analysis. YS, TN, KN and RL provided clinical input. TK and SM oversaw data collection and management. JvdM, JD, KA and LL analysed the data. CJvV drafted the manuscript. All authors critically reviewed and edited the manuscript and consented to final publication.

## Statements and declarations

### Ethical considerations

We obtained ethical approval for this study from the University of Kwazulu-Natal Biomedical Research Ethics Committee (BE646/17), the eThekwini Municipality Health Unit, the Bethesda Hospital Ethics Committee and the KwaZulu-Natal Department of Health Provincial Research Ethics Committee.

### Consent to participate

The need for informed consent was waived as de-identified, routinely collected data was analysed.

## Consent for publication

Not applicable.

### Declaration of conflicting interest

None to declare.

### Funding statement

This work was supported, in whole or in part, by the Wellcome Trust [216421/Z/19/Z] and the Gates Foundation [INV-051067]. The conclusions and opinions expressed in this work are those of the authors alone and shall not be attributed to the Foundation. Under the grant conditions of the Foundation, a Creative Commons Attribution 4.0 License has already been assigned to the Author Accepted Manuscript version that might arise from this submission.. JD is funded by the National Institute for Health and Care Research (NIHR, grant number CL-2022–13–005, to JD). The views expressed in this publication are those of the authors and not necessarily those of the NIHR, the National Health Service, or the UK Department of Health and Social Care.

### Data availability

Interested parties can request access to the data from the eThekwini Municipality Health Unit and the South African National Department of Health TB/HIV Information System (contact details obtainable upon request to JD).

## Notes

### Competing Interest Statement

The authors have declared no competing interest.

## References

1. World Health Organization. TB incidence, https://www.who.int/teams/global-tuberculosis-programme/tb-reports/global-tuberculosis-report-2023/tb-disease-burden/1-1-tb-incidence (2023, accessed 16 November 2023).

2. World Health Organization. Health data overview for the Republic of South Africa, https://data.who.int/countries/710 (2025, accessed 13 February 2025).

3. Moyo S, Ismail F, Van der Walt M, et al. Prevalence of bacteriologically confirmed pulmonary tuberculosis in South Africa, 2017-19: a multistage, cluster-based, cross-sectional survey. Lancet Infect Dis 2022. DOI: 10.1016/S1473-3099(22)00149-9.

4. Meintjes G and Maartens G. HIV-Associated Tuberculosis. N Engl J Med 2024. DOI: 10.1056/NEJMra2308181.

5. Müller M, Wandel S, Colebunders R, et al. Immune reconstitution inflammatory syndrome in patients starting antiretroviral therapy for HIV infection: a systematic review and meta-analysis. Lancet Infect Dis 2010. DOI: 10.1016/S1473-3099(10)70026-8.

6. Cerrone M, Bracchi M, Wasserman S, et al. Safety implications of combined antiretroviral and anti-tuberculosis drugs. Expert Opin Drug Saf 2020. DOI: 10.1080/14740338.2020.1694901.

7. Simbayi I, Zuma K, Zungu N, et al. South African National HIV Prevalence, Incidence, Behaviour and Communication Survey, 2017. Cape Town: HSRC Press, 2019.

8. Health Systems Trust. KwaZulu-Natal Province. In: Massyn N, Day C, Ndlovu N, et al (eds) District Health Barometer 2019/20. Durban, South Africa: Health Systems Trust, 2020.

9. Velasco M, Castilla V, Sanz J, et al. Effect of simultaneous use of highly active antiretroviral therapy on survival of HIV patients with tuberculosis. J Acquir Immune Defic Syndr 2009. DOI: 10.1097/QAI.0b013e31819367e7.

10. Abdool Karim SS, Naidoo K, Grobler A, et al. Integration of antiretroviral therapy with tuberculosis treatment. N Engl J Med 2011. DOI: 10.1056/NEJMoa1014181.

11. Blanc F, Sok T, Laureillard D, et al. Earlier versus later start of antiretroviral therapy in HIV-infected adults with tuberculosis. N Engl J Med 2011. DOI: 10.1056/NEJMoa1013911.

12. Havlir DV, Kendall MA, Ive P, et al. Timing of antiretroviral therapy for HIV-1 infection and tuberculosis. N Engl J Med 2011. DOI: 10.1056/NEJMoa1013607.

13. Uthman OA, Okwundu C, Gbenga K, et al. Optimal Timing of Antiretroviral Therapy Initiation for HIV-Infected Adults With Newly Diagnosed Pulmonary Tuberculosis: A Systematic Review and Meta-analysis. Ann Intern Med 2015. DOI: 10.7326/M14-2979.

14. Burke RM, Rickman HM, Singh V, et al. What is the optimum time to start antiretroviral therapy in people with HIV and tuberculosis coinfection? A systematic review and meta-analysis. J Int AIDS Soc 2021. DOI: 10.1002/jia2.25772.

15. Chelkeba L, Fekadu G, Tesfaye G, et al. Effects of time of initiation of antiretroviral therapy in the treatment of patients with HIV/TB co-infection: A systemic review and meta-analysis. Ann Med Surg (Lond) 2020. DOI: 10.1016/j.amsu.2020.05.004.

16. Rosen S, Maskew M, Fox MP, et al. Initiating Antiretroviral Therapy for HIV at a Patient’s First Clinic Visit: The RapIT Randomized Controlled Trial. PLoS Med 2016. DOI: 10.1371/journal.pmed.1002015.

17. Koenig SP, Dorvil N, Dévieux JG, et al. Same-day HIV testing with initiation of antiretroviral therapy versus standard care for persons living with HIV: A randomized unblinded trial. PLoS Med 2017. DOI: 10.1371/journal.pmed.1002357.

18. World Health Organization. Consolidated guidelines on HIV prevention, testing, treatment, service delivery and monitoring: Recommendations for a public health approach. Geneva, Switzerland: World Health Organization, 2021.

19. Republic of South Africa National Department of Health. ART clinical guidelines for the management of HIV in adults, pregnancy and breastfeeding, adolescents, children, infants and neonates. Pretoria, South Africa: Republic of South Africa National Department of Health, 2023.

20. Nel J, Dlamini S, Meintjes G, et al. Southern African HIV Clinicians Society guidelines for antiretroviral therapy in adults: 2020 update. South Afr J HIV Med 2020. DOI: 10.4102/sajhivmed.v21i1.1115.

21. Osler M, Hilderbrand K, Hennessey C, et al. A three-tier framework for monitoring antiretroviral therapy in high HIV burden settings. J Int AIDS Soc 2014. DOI: 10.7448/IAS.17.1.18908.

22. Republic of South Africa National Department of Health. 2019 ART clinical guidelines for the management of HIV in adults, pregnancy, adolescents, children, infants and neonates. Pretoria, South Africa: Republic of South Africa National Department of Health, 2019.

23. World Health Organization. Definitions and Reporting Framework for Tuberculosis – 2013 Revision. Geneva: World Health Organization, 2013.

24. World Health Organization. Updated recommendations on HIV prevention, infant diagnosis, antiretroviral initiation and monitoring. Geneva: World Health Organization, 2021.

25. Dorvil N, Rivera VR, Riviere C, et al. Same-day testing with initiation of antiretroviral therapy or tuberculosis treatment versus standard care for persons presenting with tuberculosis symptoms at HIV diagnosis: A randomized open-label trial from Haiti. PLoS Med 2023. DOI: 10.1371/journal.pmed.1004246.

26. Burke RM, Rickman HM, Singh V, et al. Same-day antiretroviral therapy initiation for people living with HIV who have tuberculosis symptoms: a systematic review. HIV Med 2022. DOI: 10.1111/hiv.13169.

27. Ford N, Meintjes G, Calmy A, et al. Managing Advanced HIV Disease in a Public Health Approach. Clin Infect Dis 2018. DOI: 10.1093/cid/cix1139.

28. Engelbrecht MC, Kigozi NG, Chikobvu P, et al. Unsuccessful TB treatment outcomes with a focus on HIV co-infected cases: a cross-sectional retrospective record review in a high-burdened province of South Africa. BMC Health Serv Res 2017. DOI: 10.1186/s12913-017-2406-x.

29. McNabb KC, Bergman A and Farley JE. Risk factors for poor engagement in drug-resistant TB care in South Africa: a systematic review. Public Health Action 2021. DOI: 10.5588/pha.21.0007.

30. Munro SA, Lewin SA, Smith HJ, et al. Patient adherence to tuberculosis treatment: a systematic review of qualitative research. PLoS Med 2007. DOI: 10.1371/journal.pmed.0040238.

31. Lahuerta M, Ue F, Hoffman S, et al. The problem of late ART initiation in Sub-Saharan Africa: a transient aspect of scale-up or a long-term phenomenon?. J Health Care Poor Underserved 2013. DOI: 10.1353/hpu.2013.0014.

32. INSIGHT START Study Group, Lundgren JD, Babiker AG, et al. Initiation of Antiretroviral Therapy in Early Asymptomatic HIV Infection. N Engl J Med 2015. DOI: 10.1056/NEJMoa1506816.

33. Benchimol EI, Smeeth L, Guttmann A, et al. The REporting of studies Conducted using Observational Routinely-collected health Data (RECORD) Statement. PLoS Med 2015.

34. Etoori D, Wringe A, Kabudula CW, et al. Misreporting of Patient Outcomes in the South African National HIV Treatment Database: Consequences for Programme Planning, Monitoring, and Evaluation. Front Public Health 2020. DOI: 10.3389/fpubh.2020.00100.

35. Kerschberger B, Hilderbrand K, Boulle AM, et al. The effect of complete integration of HIV and TB services on time to initiation of antiretroviral therapy: a before-after study. PLoS One 2012. DOI: 10.1371/journal.pone.0046988.

