## Supplementary Tables for "Retrospective cohort analysis of antiretroviral therapy initiation timelines and clinical outcomes in adults with HIV and TB disease in KwaZulu-Natal, South Africa"

*Supplementary Table S1: Univariable and multivariable Poisson regression models of factors associated with successful TB outcomes, among people living with HIV on TB treatment, including patients lost to follow up (N=5,548)*

| **Time to ART initiation (days)** | **Successful outcome  n/N (%)** | **Unadjusted RR**  **(95% CI)** | **Adjusted RR**  **(95% CI)** |
| --- | --- | --- | --- |
| ≤15 | 1343 / 1652 (81.3) | 1 | - |
| 16-56 | 1577 / 2011 (78.4) | 0.96 (0.93-1.00) | 0.97 (0.94-1.00) |
| 57-210 | 387 / 481 (80.5) | 0.99 (0.94-1.04) | 1.03 (0.97-1.09) |
| Not initiated within 7 months | 630 / 1404 (44.9) | **0.55 (0.52-0.59)** | **0.58 (0.54-0.63)** |

Note: Bolded values are statistically significant at p<0.05. Successful TB outcome was defined those who completed TB treatment or were cured based on sputum results.

*Supplementary Table S2: Univariable and multivariable Poisson regression models of factors associated with successful TB outcomes, among people living with HIV on TB treatment for pulmonary TB (N=4,154)*

| **Time to ART initiation (days)** | **Successful outcome  n/N (%)** | **Unadjusted RR**  **(95% CI)** | **Adjusted RR**  **(95% CI)** |
| --- | --- | --- | --- |
| ≤15 | 1233 / 1406 (87.7) | 1 | - |
| 16-56 | 1383 / 1618 (85.5) | 0.97 (0.95-1.00) | 0.98 (0.95-1.01) |
| 57-210 | 327 / 389 (84.1) | 0.96 (0.91-1.01) | 1.00 (0.95-1.05) |
| Not initiated within 7 months | 502 / 741 (67.7) | **0.77 (0.73-0.81)** | **0.81 (0.76-0.86)** |

Note: Bolded values are statistically significant at p<0.05. Successful TB outcome was defined those who completed TB treatment or were cured based on sputum results.

*Supplementary Table S3: Breakdown of outcome categories by CD4 count and time to ART initiation (N = 2,816)*

| **Time to ART initiation (days)** | **Completed n/N (%)** | **Cured  n/N (%)** | **Failed treatment n/N (%)** | **Defaulted treatment n/N (%)** | **Died  n/N (%)** |
| --- | --- | --- | --- | --- | --- |
| **Patients with CD4 <50 cells/µL (n = 575)** | | | | | |
| **≤15** | 163 (64.9) | 56 (22.3) | 1 (0.4) | 20 (8.0) | 11 (4.4) |
| **16–56** | 197 (64.2) | 73 (23.8) | 1 (0.3) | 21 (6.8) | 15 (4.9) |
| **57–210** | 8 (47.1) | 6 (35.3) | 0 (0.0) | 3 (17.6) | 0 (0.0) |
| **Patients with CD4 ≥50 cells/µL (n = 2,241)** | | | | | |
| **≤15** | 644 (63.6) | 262 (25.9) | 4 (0.4) | 86 (8.5) | 16 (1.6) |
| **16–56** | 737 (63.6) | 259 (22.4) | 9 (0.8) | 123 (10.6) | 30 (2.6) |
| **57–210** | 52 (73.2) | 5 (7.0) | 0 (0.0) | 12 (16.9) | 2 (2.8) |

*Supplementary Table S4: Univariable and multivariable Poisson regression models of factors associated with viral load suppression, among people living with HIV who initiated ART with pulmonary TB (N=2,355).*

| **Time to ART initiation (days)** | **Suppressed  n/N (%)** | **Unadjusted RR**  **(95% CI)** | **Adjusted RR**  **(95% CI)** |
| --- | --- | --- | --- |
| ≤15 | 780 / 994 (78.5) | 1 | - |
| 16-56 | 828 / 1065 (77.7) | 0.99 (0.95-1.04) | 0.99 (0.95-1.04) |
| 57-210 | 158 / 215 (73.5) | 0.94 (0.86-1.02) | 0.93 (0.84-1.02) |
| Not initiated within 7 months | 58 / 81 (71.6) | 0.91 (0.79-1.05) | 0.90 (0.78-1.05) |

Note: Bolded values are statistically significant at p<0.05. Successful TB outcome was defined those who completed TB treatment or were cured based on sputum results.
